# A Transcriptomic Severity Metric that Predicts Clinical Outcomes in Critically Ill Surgical Sepsis Patients

**DOI:** 10.1101/2021.05.28.21258014

**Authors:** Scott C. Brakenridge, Petr Starostik, Gabriella Ghita, Uros Midic, Dijoia Darden, Brittany Fenner, James Wacker, Philip A. Efron, Oliver Liesenfeld, Timothy E Sweeney, Lyle L. Moldawer

**Affiliations:** Sepsis and Critical Illness Research Center, Department of Surgery, University of Florida College of Medicine, Gainesville, Florida 32610 USA; Molecular Pathology Laboratory at Rocky Point, Department of Pathology, Immunology and Laboratory Medicine, University of Florida College of Medicine, and Clinical and Diagnostic Laboratories, Shands UF Health Science Center, Gainesville, Florida 32610 USA; Inflammatix, Inc. Burlingame, California 94010 USA

**Keywords:** biomarkers, sepsis, critical illness, genomics, proteomics, innate immunity

## Abstract

Prognostic metrics for the rapid and accurate prediction of sepsis severity that could elicit a meaningful change in clinical practice are currently lacking. Here, we evaluated a whole blood, multiplex host mRNA expression metric, Inflammatix-Severity-2 (IMX-SEV-2), for identifying septic, hospitalized patients for 30-day mortality, development of chronic critical illness (CCI), discharge disposition, and/or secondary infections.

**Methods:** This is a retrospective, validation cohort analysis of a prospectively enrolled 335 patient study with surgical sepsis treated in the surgical intensive care unit. Whole blood collected in PAXgene^®^ Blood RNA collection tubes at 24 hours post sepsis diagnosis was analyzed using a custom 29-mRNA classifier (IMX-SEV-2) in a CLIA-CAP-accredited diagnostic laboratory using the NanoString FLEX^®^ profiler.

**Results:** Among patients meeting Sepsis-3 criteria, the IMX-SEV-2 severity score was significantly better (p<0.05) at predicting secondary infections (AUROC 0.71) and adverse clinical outcomes (AUROC 0.75) than C-reactive protein (CRP), absolute lymphocyte counts (ALC), total white blood cell (WBC) count, age and Charlson comorbidity index (and better, albeit nonsignificantly, than IL-6 and APACHE II). Using multivariate logistic regression analysis, only combining the Charlson comorbidity index (AUROC 0.80) or APACHE II (AUROC 0.81) with the IMX-SEV-2 significantly improved prediction of adverse clinical outcomes, and combining with the Charlson comorbidity index for predicting 30-day mortality (AUROC 0.79).

**Conclusions:** The IMX-SEV-2 severity score was superior at predicting secondary infections and overall adverse clinical outcomes versus other common metrics. Importantly, combining a rapidly measured transcriptomic metric with clinical or physiologic indices has sufficient precision to optimize resource utilization and allow adjustments to patient management that may improve outcomes in surgical sepsis. Hospitalized patients who are septic and present with an elevated IMX-SEV2 severity score and pre-existing comorbidities would be strong candidates for clinical interventions aimed at reducing the risk of secondary infections and adverse clinical outcomes.

## INTRODUCTION

Sepsis afflicts over 1.7 million Americans annually, and accounts for over 250,000 deaths in the United States alone.^1^ Sepsis remains the most common cause of death in the intensive care unit (ICU).^2^ Approximately 60-70% of all sepsis cases are diagnosed in the emergency department (ED). 10-40% percent of total sepsis cases develop in currently hospitalized patients.^2-4^ Generally speaking, hospital-acquired sepsis, and surgical sepsis in particular, are associated with a higher frequency of septic shock and a two-fold increase in mortality compared to community acquired sepsis.^3,5^

Identifying the severity of disease and likelihood of further deterioration of surgical sepsis patients with high accuracy and rapid turnaround remains a critical unmet need for hospitalized patients, since time to intervention and antimicrobial treatment is a critical determinant of outcome.^6,7^ Current in-hospital early warning systems for sepsis (Modified Early Warning System [MEWS]) are used to alert health-care providers to the possibility of sepsis.^8^ In many cases, the decision to initiate sepsis resuscitation bundles and more intensive management is an empiric decision made by the health-care provider at the bedside, prior to documentary evidence of microbial infection, organ injury or immunological dyscrasia. Existing clinical scores such as the Acute Physiology and Chronic Health Evaluation II (APACHE II) and sequential organ failure assessment (SOFA) score require 24 hours to properly calculate, making them impractical for immediate clinical decision-making. The quick (q)SOFA assessment associated with the Sepsis-3 criteria has been recommended to identify patients at high risk of death, but its value is still controversial.^9,10^ Other less common metrics such as procalcitonin, IL-6 and monocyte distribution width (MDW) have all been promulgated as being predictive of sepsis severity, ^11,12^ although their predictive ability to either diagnose sepsis or its severity remains controversial.^13^ A rapid and clinically applicable diagnostic that can both diagnose infection and predict sepsis severity will be required to alter clinical management of the individual patient.

Here we sought to validate the ability of a novel whole-blood host immune transcriptomic metric for predicting outcome severity in an established prospective cohort of critically ill surgical/trauma patients adjudicated as being septic. This metric of sepsis severity (IMX-SEV-2) is a component of a multi-biomarker host response approach to both diagnosing infections and predicting sepsis severity, the InSep™ test (Inflammatix, Inc., Burlingame, CA, USA).^14^ Intended as a point of care diagnostic with rapid turn-around times, InSep™ will provide the clinician with actionable information for upgrading management strategy and tailor ICU resources to individual patient needs.

## MATERIALS AND METHODS

### Study Design and Subject Enrollment

This is a secondary, validation cohort analysis of a prospective, observational longitudinal study of hospitalized patients with surgical sepsis. We utilized the transcriptomic metric, IMX-SEV-2, to predict 30-day mortality when applied to samples obtained from a single-center prospective 1-year longitudinal cohort study (NCT02276417) of critically ill surgical patients on day 1 following diagnosis of sepsis. The parent study included 363 septic patients hospitalized in a 48-bed surgical intensive care unit (ICU) at a quaternary care academic hospital between January, 2015, and January, 2020. Ethics approval was obtained from the University of Florida Institutional Review Board (#201400611). Informed consent was obtained from each subject or their proxy. Detailed study cohort design and protocols utilized by the parent cohort have been published previously.^15,16^ Detailed descriptions of the inclusion and exclusion criteria are contained in the **Supplemental Methods and Results**.

All enrolled subjects underwent prospective adjudication by physician-investigators at weekly program meetings to confirm sepsis diagnosis, severity and source.^17^ Adjudicated non-septic patients were excluded from this analysis. As the parent cohort was designed prior to the publication of Sepsis-3 consensus guidelines, patients were enrolled into the sampling cohort using 2001 sepsis consensus criteria definitions.^18^ Subsequently, patients were retrospectively re-adjudicated for sepsis and septic shock using the Sepsis-3 guidelines.^19^

Hospital-acquired secondary infections were adjudicated utilizing current United States Centers for Disease Control definitions and guidelines.^20^ Discharge disposition was prospectively classified based on known associations with long-term outcomes as either ‘good’ (home with or without health care services, or rehabilitation facility) or ‘poor’ (long-term acute care facility [LTAC]), skilled nursing facility [SNF], another acute care hospital, hospice or inpatient death).

Individual clinical outcome variables included: 1) 30-day (all-cause) mortality, 2) development or absence of chronic critical illness (CCI), 3) discharge disposition, and 4) secondary infections. Inpatient clinical trajectory was defined as ‘early death’, ‘rapid recovery’ (RAP), or ‘chronic critical illness’ (CCI). Early death was defined as death within 14 days of sepsis onset. CCI was defined as an ICU length of stay (LOS) greater than or equal to 14 days with evidence of persistent organ dysfunction based upon components of the SOFA score.^21^ Hospitalized patients who died after an ICU length of stay >14 days from the index hospitalization were classified as CCI.^22^ Rapid recovery (RAP) patients were those discharged from the ICU within 14 days with resolution of organ dysfunction. Patients were defined as having an ‘adverse clinical outcome’ if they experienced a secondary infection, CCI, poor discharge disposition and/or mortality within the first 30 days.

### Sample Collection and Analyses

Blood samples were collected in PAXgene™ tubes at 24 hours following initiation of EMR-based sepsis management protocols and were stored at -80°C for subsequent bulk analysis. RNA was extracted with the RNeasy^®^Plus Micro Kit (QIAGEN, Inc., Germantown, MD, USA). The IMX-SEV-2 classifier (**Supplemental Table 1)** was quantitated from 200ng of RNA input using the 510(k)-cleared NanoString nCounter FLEX^®^profiler (NanoString, Inc., Seattle, WA. USA) according to a validated standard operating protocol in a CLIA-CAP accredited clinical/diagnostic laboratory.

Total leukocyte counts, absolute lymphocyte counts, and C-reactive protein concentrations were determined at the UF Health Clinical and Diagnostic Laboratories. Plasma interleukin-6 (IL-6) and glucagon-like peptide 1 (GLP-1) levels were determined using the Luminex MagPix^™^ platform (Austin, TX, USA).

### IMX Classifiers

We previously described the development of a 29 host-mRNA test, InSep™ (Inflammatix Inc., Burlingame, CA) that uses machine learning algorithms IMX-SEV (severity metric trained on 30-day mortality), and IMX-BVN (bacterial-viral-non-infected, for infection diagnosis)] to produce three scores for the likelihood of bacterial infection, the likelihood of viral infection and disease severity.^14,23^ Here, we utilize the second-generation versions of the neural-network-based classifiers, including IMX-BVN-2 and IMX-SEV-2. Each score is reported both as a continuous variable and stratified into pre-set ‘risk bands’ to meet clinically actionable performance thresholds.^14^ For IMX-SEV-2, the score is broken into three categories: “likely”, “possible” and “unlikely” of 30-day mortality, based on prior published work (see **Supplemental Figure 1**).^14,23^

### Statistical Analyses

Descriptive data are presented as frequency and percentage, mean and standard deviation, or median and 25th/75th percentiles. Fisher’s exact test and the Kruskal–Wallis test were used for comparison of categorical and continuous variables, respectively. Correlations among continuous and discrete variables were determined using Spearman’s test. Area under the receiver operating curve values (AUROC) and Hosmer–Lemeshow goodness-of-fit test were used to assess model discrimination and fit. The DeLong test was used to compare differences among receiver operating curves. False discovery correction for multiple comparisons was performed using the Benjamini–Hochberg procedure. The continuous Net Reclassification Index (cNRI)^24^ was employed to measure the improvement in prediction performance gained by adding a clinical metric to the IMX-SEV-2 score. The cNRI was calculated using R (version 4.0.5, https://www.r-project.org/). All remaining analyses were performed using SAS (v.9.4, Cary, NC, USA). All significance tests were two-sided, with a p-value ≤0.05 considered statistically significant.

## RESULTS

The parent study was enrolled a total of 363 sepsis patients that met 2001 consensus sepsis criteria and provided informed consent. Of these 363 patients, remaining RNA samples were available for this analysis in 333 subjects at 24 hours (± 6 hours), and two samples were obtained from additional subjects at less than 12 hours from the initiation of the sepsis management protocols. Of the 335 patients in this “overall” cohort, 316 subsequently met the criteria for sepsis or septic shock per Sepsis-3 criteria.^19^ (**Supplemental Table 2**).

Both cohorts represented an older population (median, 62 years) with a significant burden of pre-existing comorbidities (median Charlson comorbidity index, 3). Approximately 25% of Sepsis-3 patients met shock criteria; nearly 60% presented with or developed acute kidney injury, and nearly half progressed to multiple organ failure (MOF). More than half of the patients had either intra-abdominal or surgical site infection, while approximately 28% had either pneumonia or urinary tract infections. Secondary infections were also common in this Sepsis-3 cohort, with an overall rate of 2.1 infections per 100 person hospital days (**Supplemental Table 3**). Thirty patients (9.5%) died and 194 patients (61.4%) had what was defined as an adverse clinical outcome (**Supplemental Figure 2)**.

### Predicting Clinical Outcomes

The prognostic ability of the IMX-SEV-2 severity metric was analyzed both as a continuous variable and as likelihood distributions based on predetermined thresholds (**Supplemental Figure 1**). For patients meeting Sepsis-3 criteria, the IMX-SEV-2 metric not only predicted 30-day mortality, but also predicted development of CCI, discharge disposition, and incidence of secondary infections (**Table 1). Table 2** presents the actual test characteristics of the IMX-SEV-2 metric when presented as likelihood distributions. Sensitivity and specificity were calculated by combining the “possible” group with the “likely” and “unlikely” groups, respectively.

**Table 1.**
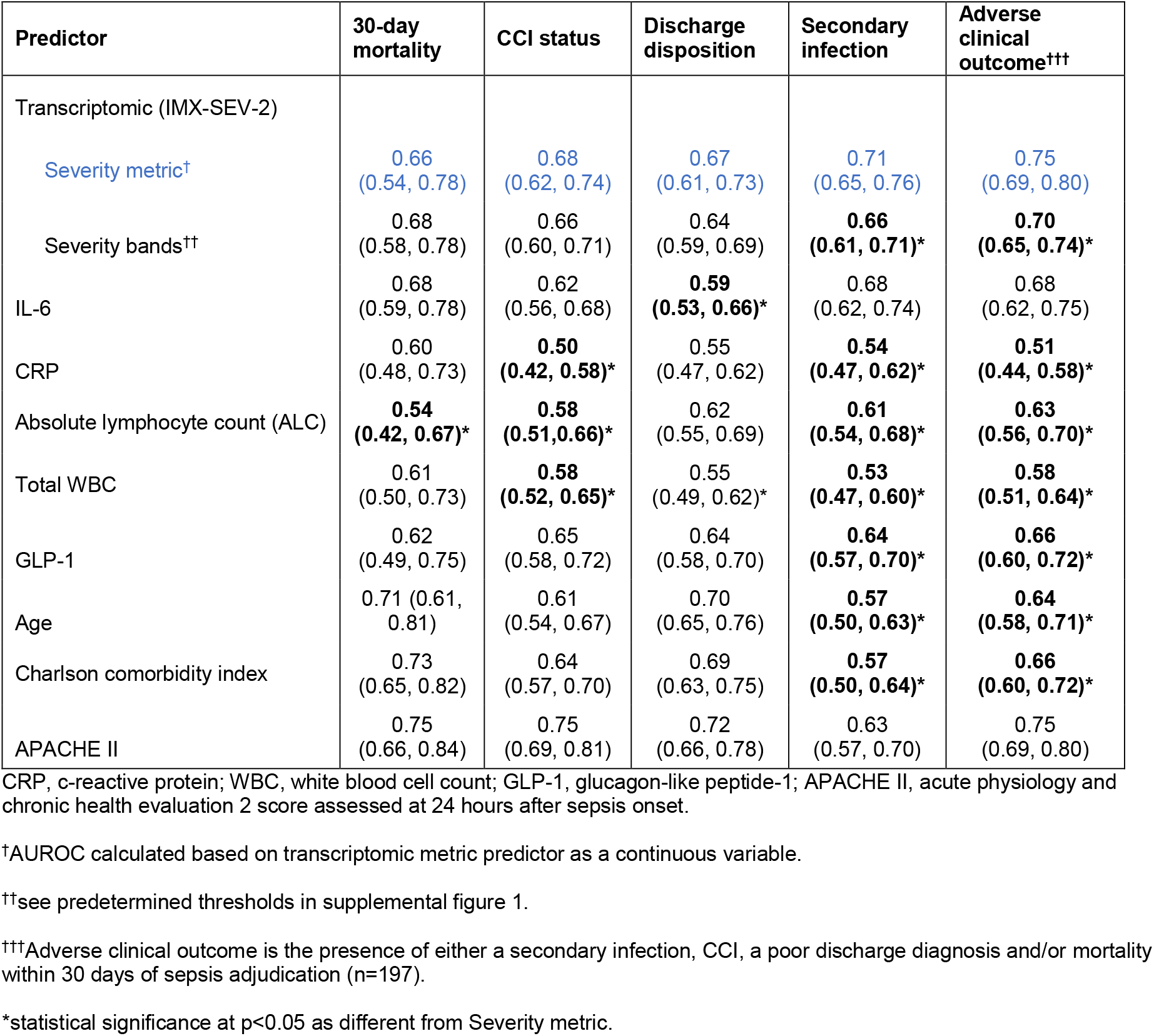
Area Under the Receiver-Operator Curve (AUROC) for Outcome Variables. Values were obtained in patients meeting Sepsis-3 criteria for 30 day mortality, development of chronic critical illness (CCI), discharge disposition (“Good” vs. “Poor”), presence of secondary infection or an adverse clinical outcome.

| Predictor | 30-day mortality | CCI status | Discharge disposition | Secondary infection | Adverse clinical outcome<sup>†††</sup> |
| --- | --- | --- | --- | --- | --- |
| Transcriptomic (IMX-SEV-2) |  |  |  |  |  |
| Severity metric<sup>†</sup> | 0.66<br>(0.54, 0.78) | 0.68<br>(0.62, 0.74) | 0.67<br>(0.61, 0.73) | 0.71<br>(0.65, 0.76) | 0.75<br>(0.69, 0.80) |
| Severity bands<sup>††</sup> | 0.68<br>(0.58, 0.78) | 0.66<br>(0.60, 0.71) | 0.64<br>(0.59, 0.69) | **0.66**<br>**(0.61, 0.71)\*** | **0.70**<br>**(0.65, 0.74)\*** |
| IL-6 | 0.68<br>(0.59, 0.78) | 0.62<br>(0.56, 0.68) | **0.59**<br>**(0.53, 0.66)\*** | 0.68<br>(0.62, 0.74) | 0.68<br>(0.62, 0.75) |
| CRP | 0.60<br>(0.48, 0.73) | **0.50**<br>**(0.42, 0.58)\*** | 0.55<br>(0.47, 0.62) | **0.54**<br>**(0.47, 0.62)\*** | **0.51**<br>**(0.44, 0.58)\*** |
| Absolute lymphocyte count (ALC) | **0.54**<br>**(0.42, 0.67)\*** | **0.58**<br>**(0.51, 0.66)\*** | 0.62<br>(0.55, 0.69) | **0.61**<br>**(0.54, 0.68)\*** | **0.63**<br>**(0.56, 0.70)\*** |
| Total WBC | 0.61<br>(0.50, 0.73) | **0.58**<br>**(0.52, 0.65)\*** | 0.55<br>(0.49, 0.62)* | **0.53**<br>**(0.47, 0.60)\*** | **0.58**<br>**(0.51, 0.64)\*** |
| GLP-1 | 0.62<br>(0.49, 0.75) | 0.65<br>(0.58, 0.72) | 0.64<br>(0.58, 0.70) | **0.64**<br>**(0.57, 0.70)\*** | **0.66**<br>**(0.60, 0.72)\*** |
| Age | 0.71 (0.61, 0.81) | 0.61<br>(0.54, 0.67) | 0.70<br>(0.65, 0.76) | **0.57**<br>**(0.50, 0.63)\*** | **0.64**<br>**(0.58, 0.71)\*** |
| Charlson comorbidity index | 0.73<br>(0.65, 0.82) | 0.64<br>(0.57, 0.70) | 0.69<br>(0.63, 0.75) | **0.57**<br>**(0.50, 0.64)\*** | **0.66**<br>**(0.60, 0.72)\*** |
| APACHE II | 0.75<br>(0.66, 0.84) | 0.75<br>(0.69, 0.81) | 0.72<br>(0.66, 0.78) | 0.63<br>(0.57, 0.70) | 0.75<br>(0.69, 0.80) |

CRP, c-reactive protein; WBC, white blood cell count; GLP-1, glucagon-like peptide-1; APACHE II, acute physiology and chronic health evaluation 2 score assessed at 24 hours after sepsis onset.
<sup>†</sup>AUROC calculated based on transcriptomic metric predictor as a continuous variable.
<sup>††</sup>see predetermined thresholds in supplemental figure 1.
<sup>†††</sup>Adverse clinical outcome is the presence of either a secondary infection, CCI, a poor discharge diagnosis and/or mortality within 30 days of sepsis adjudication (n=197).
\*statistical significance at p<0.05 as different from Severity metric.

**Table 2.** Point Estimates of Specificity and Sensitivity for the Primary and Secondary Outcomes.

**A. 30 Day Mortality**
|  | Dead at 30d | Alive at 30d | Sensitivity | Specificity | Likelihood Ratio |
| --- | --- | --- | --- | --- | --- |
| Likely | 17 | 57 | 0.567 | 0.792 | 2.72 |
| Possible | 11 | 186 |  |  |  |
| Unlikely | 2 | 31 | 0.933 | 0.133 | 0.59 |

**B. Development of Chronic Critical Illness**
|  | CCI or Early Death | Rapid Recovery | Sensitivity | Specificity | Likelihood Ratio |
| --- | --- | --- | --- | --- | --- |
| Likely | 51 | 24 | 0.392 | 0.871 | 3.04 |
| Possible | 75 | 133 |  |  |  |
| Unlikely | 4 | 29 | 0.969 | 0.156 | 0.20 |

**C. Discharge Status**
|  | Poor | Good | Sensitivity | Specificity | Likelihood Ratio |
| --- | --- | --- | --- | --- | --- |
| Likely | 50 | 25 | 0.355 | 0.857 | 2.48 |
| Possible | 86 | 122 |  |  |  |
| Unlikely | 5 | 28 | 0.965 | 0.160 | 0.22 |

**D. Secondary Infection**
|  | Secondary Infection | No Secondary Infection | Sensitivity | Specificity | Likelihood Ratio |
| --- | --- | --- | --- | --- | --- |
| Likely | 42 | 33 | 0.382 | 0.840 | 2.38 |
| Possible | 68 | 140 |  |  |  |
| Unlikely | 0 | 33 | 1 | 0.160 | 0 |

**E. Adverse Clinical Outcome**
|  | Adverse Outcome | No Adverse Outcome | Sensitivity | Specificity | Likelihood Ratio |
| --- | --- | --- | --- | --- | --- |
| Likely | 67 | 8 | 0.345 | 0.934 | 5.27 |
| Possible | 121 | 87 |  |  |  |
| Unlikely | 6 | 27 | 0.969 | 0.221 | 0.14 |

*Values represent the number of subjects in each likelihood distribution category. Sensitivity and specificity were calculated by collapsing the likely and possible, and possible and unlikely categories, respectively.*

As shown in **Table 1** and **Figure 1**, the continuous IMX-SEV-2 severity metric significantly better predicted the incidence of secondary infections than plasma CRP, plasma GLP-1, ALC, total WBC count, age and Charlson comorbidity index (all p<0.05). Additionally, the IMX-SEV-2 metric showed equivalency to predicting 30-day mortality and discharge disposition to other physiological markers, and significantly improved (p<0.05) predictive ability when compared to ALC for 30-day mortality, and IL-6 for discharge disposition (**Table 1**). Finally, the IMX-SEV-2 severity metric better predicted (p<0.05) the composite adverse clinical outcome than all of the other metrics other than IL-6 and APACHE II score.

**Figure 1.**
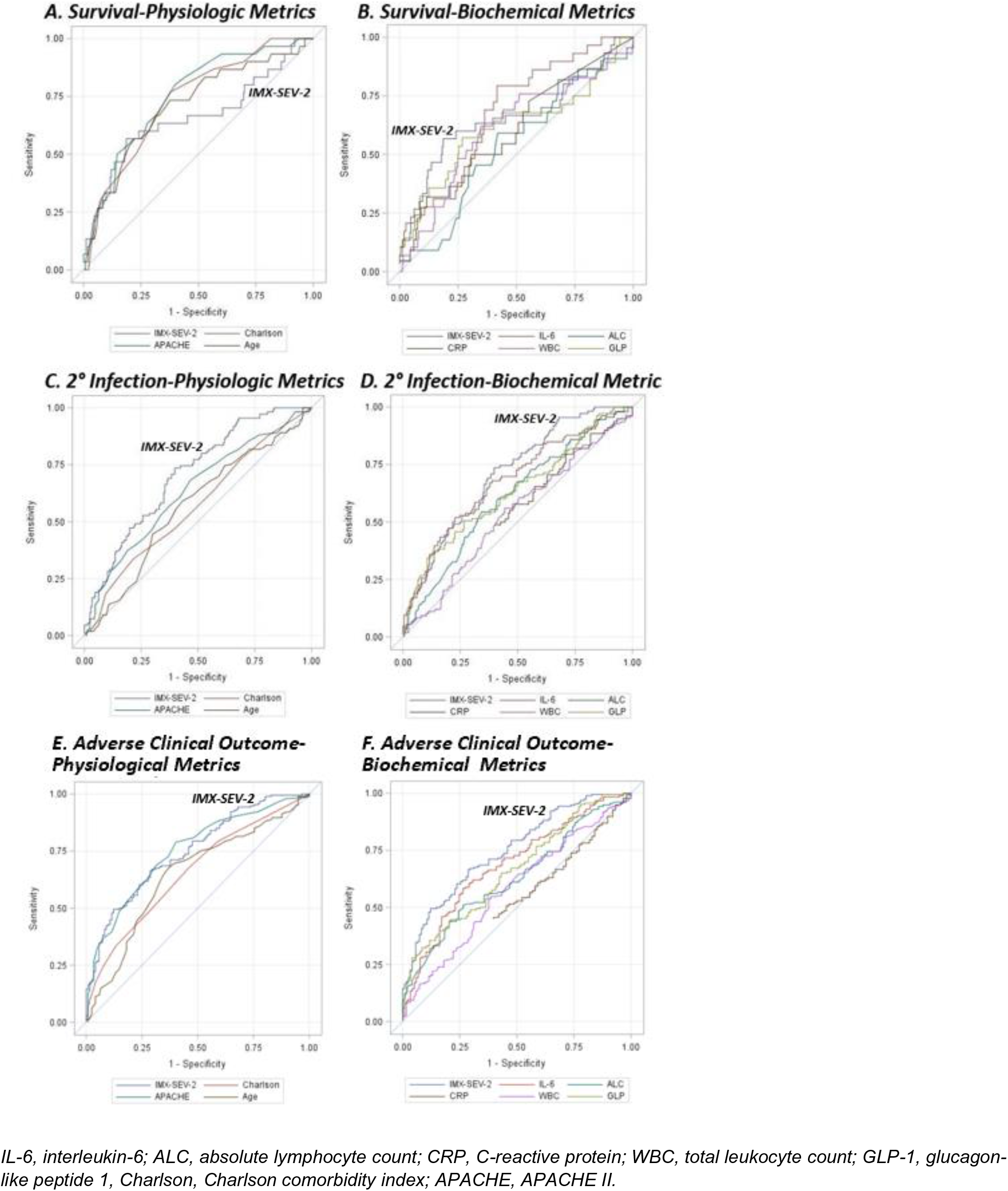
AUROC performance curves for 30 day mortality (panel A and B) and incidence of secondary infection (panel C and D) using the continuous IMX-SEV-2 metric compared to other physiologic (panel A and C) and biochemical markers (panel B and D).

### Correlation between Severity Index and Other Metrics

Spearman correlation coefficients were determined among the IMX-SEV-2 severity index and other biochemical and physiologic indices. The severity index was most closely correlated (in descending order) with plasma IL-6 (ρ =0.43), GLP-1 (ρ =0.44), and APACHE II (ρ =0.34, all p<0.001). In contrast, the IMX-SEV-2 metric was not correlated with either Charlson comorbidity index or CRP levels. Weak, but significant, associations were seen with age, total WBC, and absolute lymphocyte count (all p<0.01; **Supplemental Table 4**).

### IMX-SEV-2 in Combination with Clinical Indices

Because of the strong correlations between the IMX-SEV-2 severity metric with IL-6, GLP-1 and APACHE II, but not with the Charlson comorbidity score or age, an unbiased, multivariate logistic regression analysis was conducted with all of the biochemical and clinical metrics. Only the IMX-SEV-2 metric and the Charlson comorbidity index were selected as significant and independent predictors of 30-day mortality, with an enhanced composite AUROC of 0.79 (**Table 3A, Figure 2**). Similarly, combining the IMX-SEV-2 severity metric with the APACHE II score or Charlson comorbidity index for predicting adverse clinical outcomes increased the AUROC to 0.81 (**Table 3B**) and 0.80, respectively.

**Table 3.** Combining IMX-SEV-2 Metric with Clinical and Biochemical Indices to Predict 30 Day Mortality (Panel A) and Adverse Clinical Outcomes (Panel B).

| Multivariate model | Odds Ratio (95% CI) | p |
| --- | --- | --- |
| IMX-SEV-2 Severity metric (increase of 0.1) | 1.92 (1.40, 2.64) | <0.0001 |
| Charlson comorbidity index | 1.36 (1.17, 1.57) | <0.0001 |
| AUROC (95% CI) = 0.79 (0.71, 0.87) |  |  |

| Multivariate model | Odds Ratio (95% CI) | P |
| --- | --- | --- |
| IMX-SEV-2 Severity metric (increase of 0.1) | 4.42 (2.14, 9.17) | <0.0001 |
| APACHE II | 1.11 (1.05, 1.18) | 0.0003 |
| AUROC (95% CI) = 0.80 (0.74, 0.87) |  |  |

**Figure 2.**
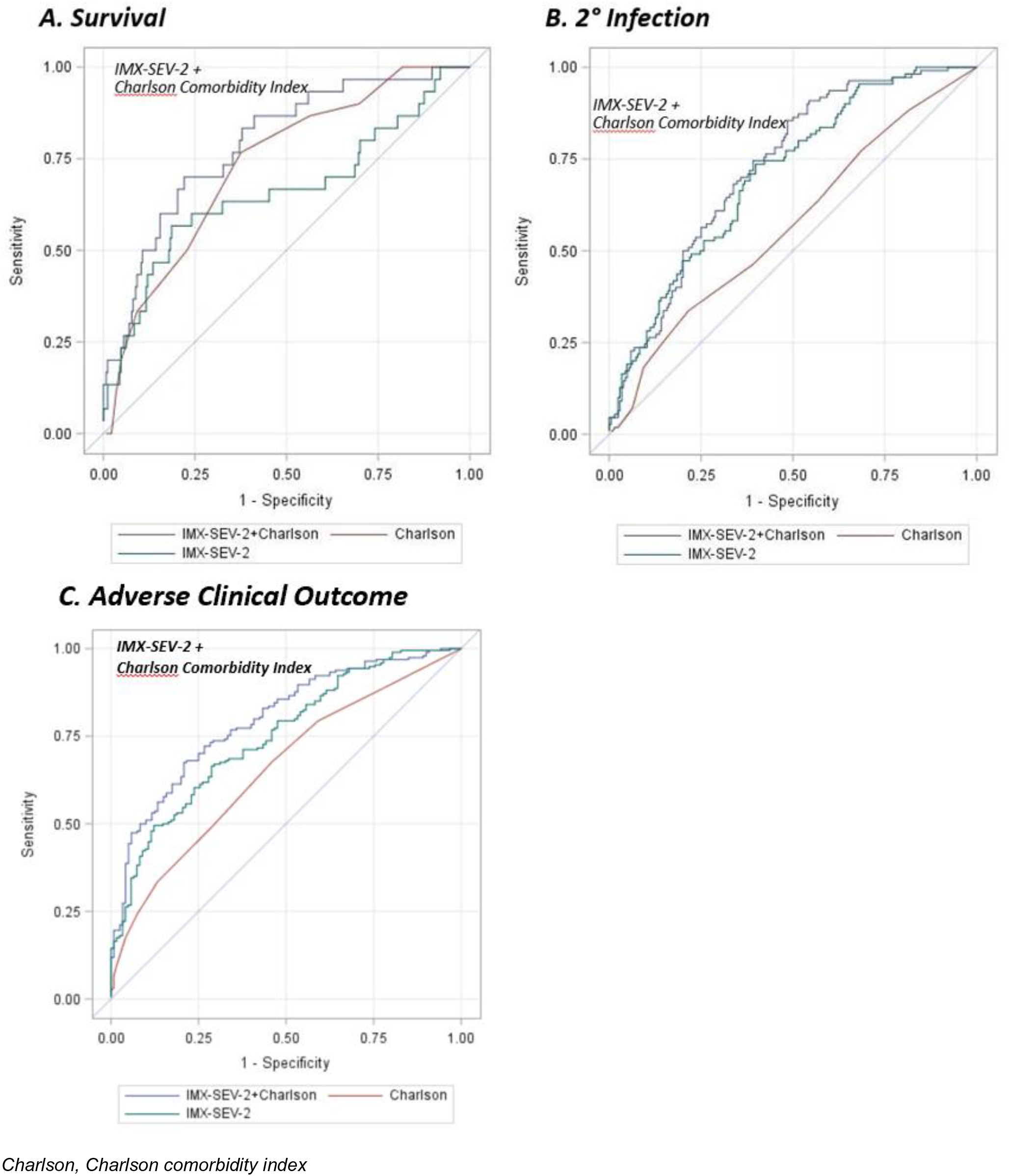
AUROC performance curves for 30 day mortality (panel A) and incidence of secondary infection (panel B) using the continuous IMX-SEV-2 metric combined with the Charlson comorbidity index compared to each alone.

A more clinically relevant measure of the usefulness of combining a clinical index with the IMX-SEV-2 metric employs the cNRI ^24^ although its use remains controversial.^25^ Using the cNRI to calculate the benefit of adding the IMX-SEV-2 metric to the Charlson comorbidity index improved the reclassification score (0.79, 95% bootstrap CI: 0.20-0.99; p<0.05) suggesting a 79% improvement in the reclassification rate for 30-day mortality when the two metrics were combined. In addition, even though IMX-SEV-2 metric was strongly correlated with the APACHE II score, adding the predictive ability of the IMX-SEV-2 metric to the APACHE II score for 30-day mortality generated a cNRI metric (0.32, CI: 0.13-0.45, p<0.05), indicative of a 32% reclassification rate.

Similar improvements in the reclassification score were seen for predicting adverse outcomes when adding the IMX-SEV-2 metric to the Charlson comorbidity index and APACHE II, respectively (cNRI, 0.71 and 0.69, both p<0.05).

### Infection diagnosis with IMX-BVN-2

As the cohort contained only infected patients with clinically adjudicated bacterial sepsis who had received antibiotics in the past 24 hours, the likelihood ratios for the bacterial and viral infection metrics (IMX-BVN-2) are less relevant, and therefore, have been relegated to **Supplemental Tables 5 and 6**.

## DISCUSSION

Surgical sepsis remains one of the most common and expensive hospital complications, and has been a major target for health care quality improvements. Institution of early warning systems that can identify patients at risk of decompensation are now routinely used in academic health centers. The clinical challenge in sepsis with these early warning systems remains the timely diagnosis of microbial infection and the recognition of severity of the consequential inflammatory insult since both affect initial management and appropriate selection of intensity of monitoring and care.

The InSep™ acute infection and sepsis test interprets 29 host-immune mRNAs together with integrating classifiers to determine the likelihood of a bacterial or viral infection and the severity of the condition.^14^ InSep™ is currently being developed as a sample-to-answer, cartridge-based point of care test with a 30-minute turnaround time, but in this study the IMX-SEV-2 classifiers were run in a CLIA-certified, CAP-accredited clinical diagnostic platform with an overnight turn-around time. Shorter analytical time and point of care assessment will increase its diagnostic value, especially versus physiologic metrics or clinical scores that cannot be measured until 12-24 hours, improving health care decisions that meet the needs of the rapidly evolving septic patient.

Importantly, the IMX-SEV-2 severity neural-network classifier was trained on multiple transcriptomic databases from patients with both community-as well as hospital-acquired sepsis.^23^ Prospective validation of the IMX-SEV-2 metric for assessing the likelihood of 30-day mortality was recently obtained in a community-acquired sepsis cohort with AUROC scores exceeding 0.80.^26,27^

When utilizing the predetermined IMX-SEV-2 severity band categories (**Supplemental Figure 1**), the AUROC for predicting 30-day mortality in surgical sepsis was similar to other laboratory metrics and clinical indices. However, when considered as a continuous variable, the metric was significantly better at predicting both secondary infections (AUROC 0.71) and an overall adverse clinical outcome (AUROC 0.75) versus all the other metrics with the exception of IL-6 and APACHE II scores, (**Table 1**). It is important to note that APACHE II is both a measure of physiologic derangement that is collinear to IMX-SEV-2, and is a composite score including age. Additionally, it is retrospectively calculated utilizing the most aberrant parameters measured within the first 24 hours of ICU admission. Thus, APACHE II is not a practically usable point of care assessment within the first hours after suspicion of sepsis diagnosis. The observation that the IMX-SEV-2 metric was at least equivalent, and more often better than all other metrics at predicting a variety of clinical outcomes in this complicated hospitalized surgical sepsis population is remarkable.

It was not surprising that the IMX-SEV-2 severity metric was correlated with other measures of the inflammatory response (IL-6) or physiological derangement (APACHE II) (**Supplemental Table 4**), since the IMX-SEV-2 severity metric incorporates transcriptomic changes that quantitate the magnitude of the host inflammatory response. Interestingly, the metric showed no significant correlation with the Charlson co-morbidity score despite the fact that the 30-day mortality predictive properties were similar (**Table 1**). This suggests that the two metrics are independently predicting survival. This led us to examine whether combining the IMX-SEV-2 metric with clinical indices could improve the overall predictive power. Importantly, when the Charlson index was combined with the IMX-SEV-2 metric, the AUROC improved significantly to 0.79 and 0.81 for 30-day mortality and adverse clinical outcomes, respectively. The cNRI suggested that the improvement in 30-day mortality and adverse clinical outcome predictive ability were achieved by an increase in identifying true positives (sensitivity).

An important question is whether (and how) rapid and improved prognostication might make a difference in clinical practice. Metrics like AUROC and cNRI aren’t useful to a bedside clinician; but IMX-SEV-2 has pre-set interpretation bands tested here with high accuracy (low-severity group 97% sensitive and high-severity group 93% specific for combined poor outcomes). Critical care medicine is an art and a science, and one of the most common questions is whether to escalate organ support in a patient not clearly recovering. We posit that a patient with a ‘low-severity’ result could be considered for a ‘stay the course’ approach, while a ‘high-severity’ result may benefit from earlier and more aggressive monitoring or organ support interventions. Additionally, the demonstrated high sensitivity of IMX-SEV-2 to rule out 30-day mortality or adverse clinical outcomes could make it valuable triage tool in critical care resource-limited environments or pandemic/disaster scenarios. Of course, this hypothesis requires interventional testing before implementation.

There are, several caveats that need to be considered. A significant proportion of the patients (n=123/316, 39%) were hospitalized for another medical condition, and sepsis was hospital-acquired and/or secondary to some injurious process. Thus, these patients were likely to have had an ongoing inflammatory process prior to the sepsis event. How that pre-existing inflammatory process influenced the subsequent host response to the sepsis is unknown, but is clearly a complicating factor. With that said, however, the findings emphasize the generalizability of the test and the power of using large datasets to generate transcriptomic signatures.^28,29^

Additionally, the metric was studied here in hospitalized patients already adjudicated to having sepsis. Blood samples were collected not at the time of sepsis suspicion, but between 12 and 24 hours after the presumption of sepsis and onset of treatment bundles, including initiation of broad-spectrum antibiotics. The primary use of IMX-SEV-2 in hospitalized patients will be in individuals *suspected* of having sepsis based on their early-warning system scores, prior to the initiation of sepsis treatment bundles. A prospective validation of the metric in hospitalized patients suspected of sepsis is currently underway.

## CONCLUSIONS

In this validation cohort of clinically-adjudicated surgical sepsis patients who had received initial sepsis management, a novel host-response metric (IMX-SEV-2) measured at 24 hours better predicted secondary infections and adverse clinical outcomes. Furthermore, combining the IMX-SEV-2 severity metric at 24 hours after initiation of sepsis treatment with an assessment of comorbidities could predict adverse clinical outcomes with the greatest precision. Such information could alter management and impact survival in hospitalized patients with surgical sepsis. Prognostic enrichment would also be critical in potential subject selection for future precision medicine trials in sepsis.

## Data Availability

The data from this study is available upon contact of the primary investigators and Inflammatix, Inc.

## CONTRIBUTIONS

SB, OL, TS, PL and LLM conceived and designed the study. SB, OL, TS, PAE and LLM wrote and edited the manuscript. PS developed and validated the analytical methodologies for IMX-SEV-2, under CLIA-CAP accreditation. JW, UM and GG developed the statistical analysis plan and optimized the IMX-SEV-2 likelihood bands, BF and DD edited the manuscript, and composed the figures and tables.

The authors wish to acknowledge the efforts of the Sepsis and Critical Illness Research Center research nursing staff, Jennifer Lanz, Ruth Davis, Ashley McCray and Jillianne Brakenridge, and research laboratory staff, Ricardo Ungaro, Marvin Dirain, Dina Nacionales, and Jaimar Rincon for the conduct of the parent clinical study. The authors also recognize the efforts of Mei Zhang from the Molecular Pathology Laboratory at Rocky Point for the Nanostring analyses for the IMX-SEV-2 measurements.

## SUPPLEMENTAL DIGITIAL CONTENT

### Supplemental Methods

Samples were collected and processed from 335 newly adjudicated surgical sepsis patients, and blood samples were collected at <12 (n=2) and 24 hours (n=333). Upon presumption of sepsis (to be adjudicated at a later point of time), patients were transferred to the surgical ICU (if already not there) where they received implementation of evidence-based sepsis management protocols, including fluid resuscitation, broad spectrum antibiotics, and inotropes (as required). Blood lactate levels were measured every four hours. The estimated time from MEWS screening/warning to presumption of sepsis and initiation of sepsis management protocols was generally less than 4 hours.

Overall cohort inclusion criteria included: 1) age ≥18 years; 2) clinical diagnosis of sepsis as defined by 2001 consensus guidelines; and 3) entrance into the electronic medical record-(EMR) based sepsis clinical management protocol.^30^ Exclusion criteria included of any of the following: 1) refractory shock (death <24 hours from sepsis protocol initiation) or inability to achieve source control (e.g., total bowel ischemic necrosis); 2) pre-admission expected lifespan <3 months; 3) patient/proxy not committed to aggressive management; 4) severe congestive heart failure (CHF) (NYHA Class IV); 5) Child-Pugh Class C liver disease or pre-liver transplant; 6) known HIV with CD4^+^ count <200 cells/mm^3^; 7) patients receiving chronic corticosteroids or immunosuppressive agents, including organ transplant recipients; 8) pregnancy; 9) institutionalized patients; 10) inability to obtain informed consent within 96 hours of enrollment; 11) chemotherapy or radiotherapy within 30 days; 12) severe traumatic brain injury (TBI); and 13) spinal cord injury (SCI) resulting in permanent sensory and/or motor deficits.

The IMX-BVN-2 and IMX-SEV-2 algorithms were evaluated on 335 subjects in whom whole blood RNA collections were available. Of those, 333 samples were obtained the following morning after diagnosis of sepsis (T1 = 24 hours) and institution of treatment bundles. Two samples were not available at 24 hours and were replaced with samples obtained at less than 12 hours post institution of treatment bundles. 289 samples had been stored in the original PAXgene collection tubes, while 46 samples were stored at -80°C as isolated total cellular RNA. The IMX-SEV-2™ metric was analyzed on a NanoString nCounter FLEX^®^using the analytical protocol, completed July 2020, using a standard 18-hour incubation period. The analyses were conducted at the University of Florida Department of Pathology Molecular Pathology Laboratory at the Rocky Point Laboratory of UF Health Shands Hospital. The Rocky Point Laboratory is CLIA-certified, CAP-accredited, and the samples were all run by a single ASCP-certified medical technologist. Quality control and quality assurances were provided according to the analytical protocol and verified by a College of Medicine faculty certified by the American Board of Pathology.

**Supplemental Table 1.** List of 29 genes used in IMX-BVN-2 and IMX-SEV-2. The scores use individual levels of mRNAs originally described in several sepsis diagnostic/prognostic modules. The mRNA modules are: (1) infection-up: *CEACAM1, ZDHHC19, C9orf95, GNA15, BATF, C3AR1;* (2) infection-down: *KIAA1370, TGFBI, MTCH1, RPGRIP1, HLA-DPB1;* (3) bacterial-viral-up: *HK3, TNIP1, GPAA1, CTSB;* (4) bacterial-viral-down: *IFI27, JUP, LAX1;* (5) mortality-up: *DEFA4, CD163, RGS1, PER1, HIF1A, SEPP1, C11orf74, CIT;* and (6) mortality-down: *LY86, TST, KCNJ2*.

Blood samples collected in PAXgene™ Blood RNA tubes had total RNA isolated as described in the primary manuscript, and RNA expression levels from 29 genes determined (Department of Pathology Molecular Pathology Laboratory at Rocky Point, Shands UF Health) (**Supplemental Table 1**). Completely blinded raw data from the NanoString nCounter FLEX^®^were transferred electronically to Inflammatix for generation of the IMX-BVN-2 and IMX-SEV-2 scores, as previously described.^23,27,31^ IMX-SEV-2 was derived from transcriptomic datasets obtained from 33 independent cohorts with a total of 3288 patients, and was validated in 348 patients, where it had an AUROC of 0.840 for predicting 30-day mortality.^28^

Diagnostic thresholds were set based on prespecified criteria so that the bacterial, viral and severity scores would be separated into discrete interpretation bands each (**Supplemental Figure 1**). The thresholds were originally set in training data targeting likelihood ratios (LRs) of 0.05 in the low bands, and 10 in the upper bands. The selected cutoffs were 0.096, 0.317, and 0.537 for the bacterial score, and 0.075, 0.288, and 0.502 for the viral score.

**Supplemental Figure 1.**
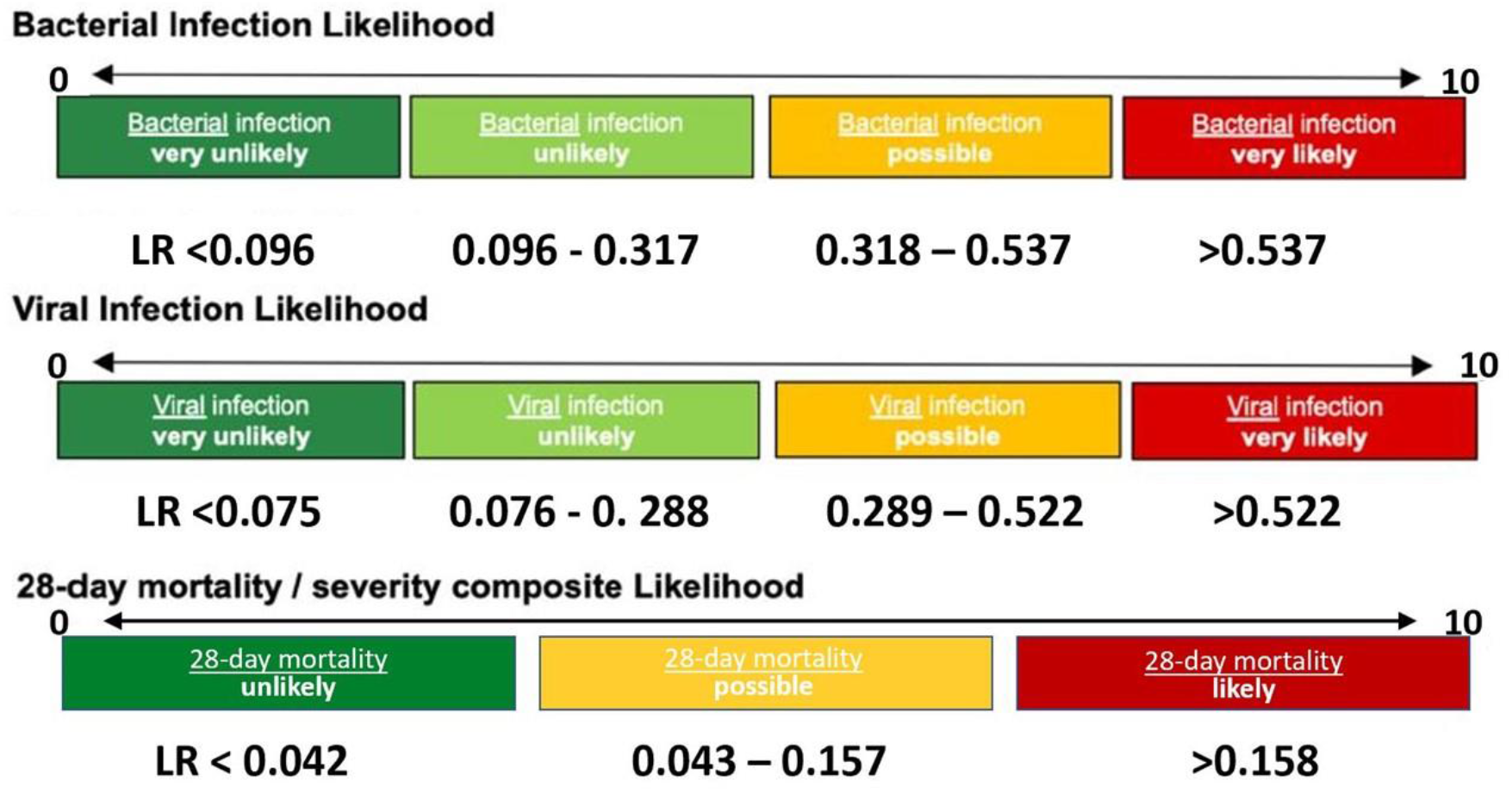
Diagnostic thresholds for likelihood of a bacterial or viral infection, or 28 (30) day mortality.

The primary clinical outcome variable was 30-day (all-cause) mortality. Secondary clinical outcome variables included: 1) development or absence of chronic critical illness (CCI), 2) hospital discharge disposition, 3) presence or absence of secondary infections and 4) an overall adverse clinical outcome. Inpatient clinical trajectory was defined as ‘early death’, ‘rapid recovery’ (RAP), or ‘chronic critical illness’ (CCI). Early death was defined as death within 14 days of sepsis onset. CCI was defined as an ICU length of stay (LOS) greater than or equal to 14 days with evidence of persistent organ dysfunction based upon components of the SOFA score.^21^ Hospitalized patients who died after an ICU length of stay >14 days from the index hospitalization were classified as CCI.^22^ Rapid recovery (RAP) patients were those discharged from the ICU within 14 days with resolution of organ dysfunction.

**Supplemental Figure 2.**
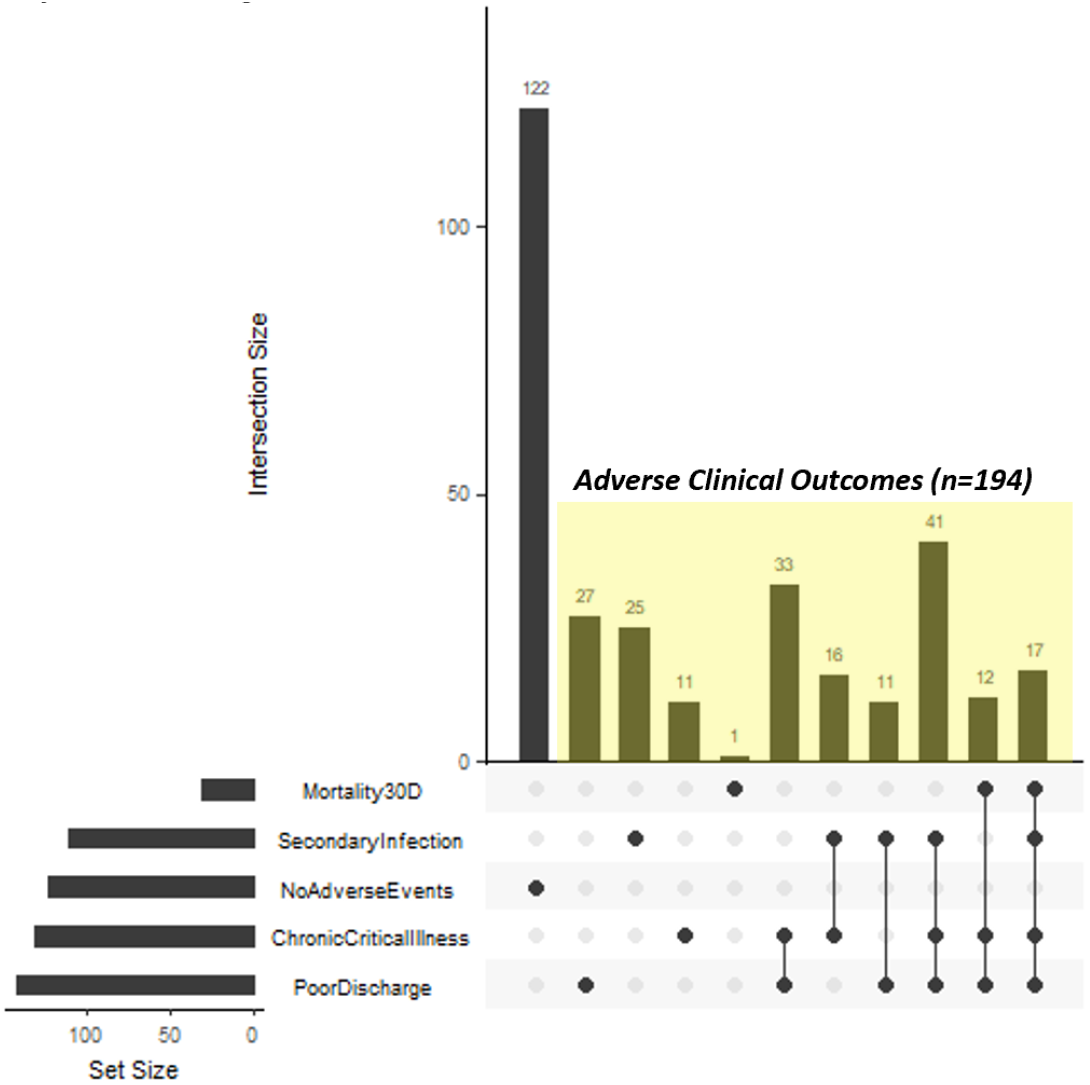
Outcome overlaps among Sepsis-3 patients. Subjects were assigned to different categories based on whether they exhibited one or more adverse clinical outcomes. The definition of an adverse clinical outcome was based on subjects exhibiting one or more of these clinical outcomes.

**Supplemental Figure 2** presents the distribution of Sepsis-3 patients with different clinical outcomes. Patients were defined as having an ‘adverse clinical outcome’ if they experienced a secondary infection, CCI, poor discharge disposition and/or mortality within the first 30 days. Outcomes overlaps were plotted with UpSetR™ for R software (https://doi.org/10.1093/bioinformatics/btx364). One hundred, twenty four Sepsis-3 patients had no adverse clinical outcomes, while 192 patients experienced one or more adverse clinical events. One hundred and three patients developed chronic critical illness (CCI) and had either an adverse discharge disposition or death.

### Supplemental Results

**Supplemental Table 2.**
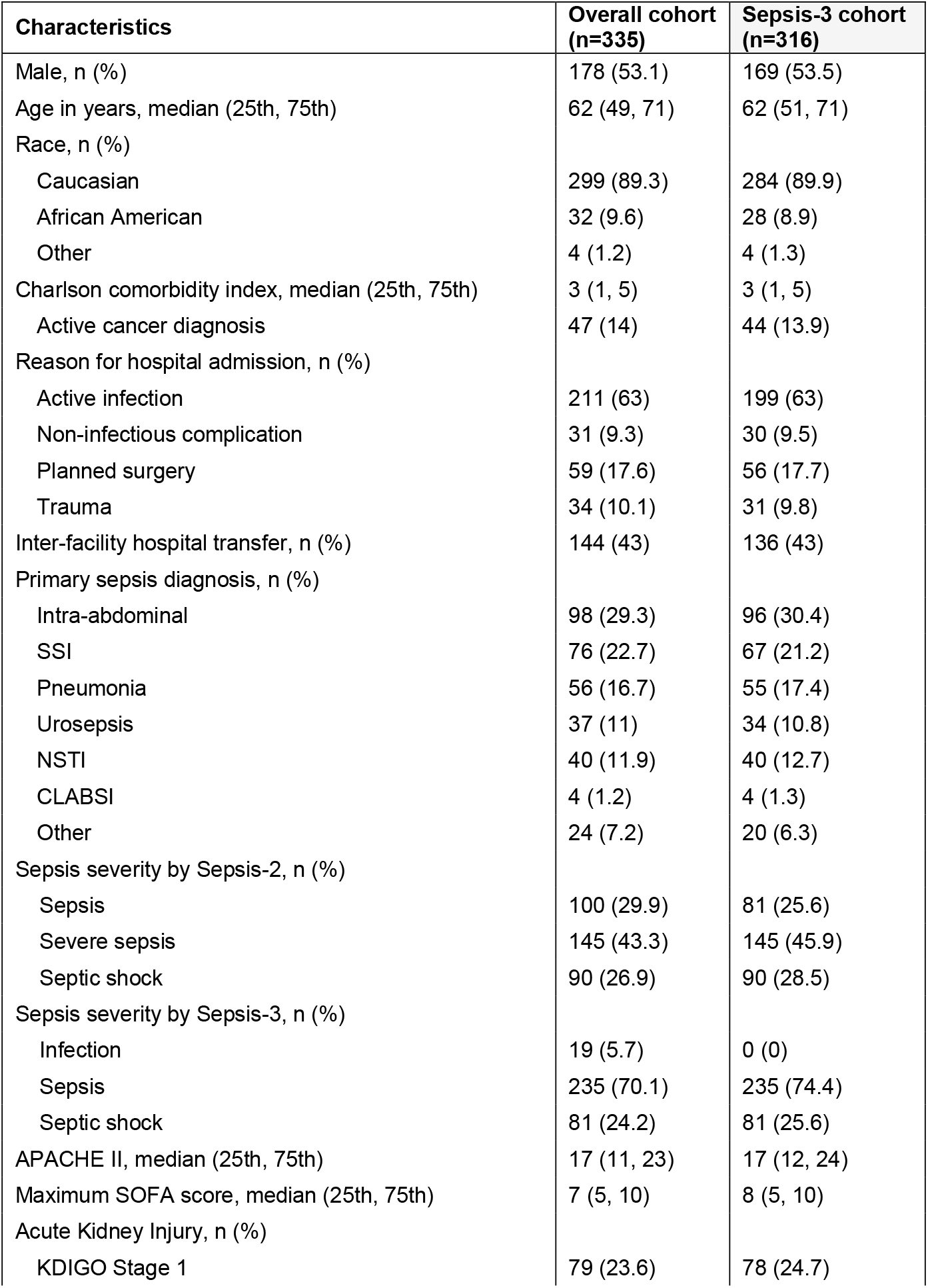
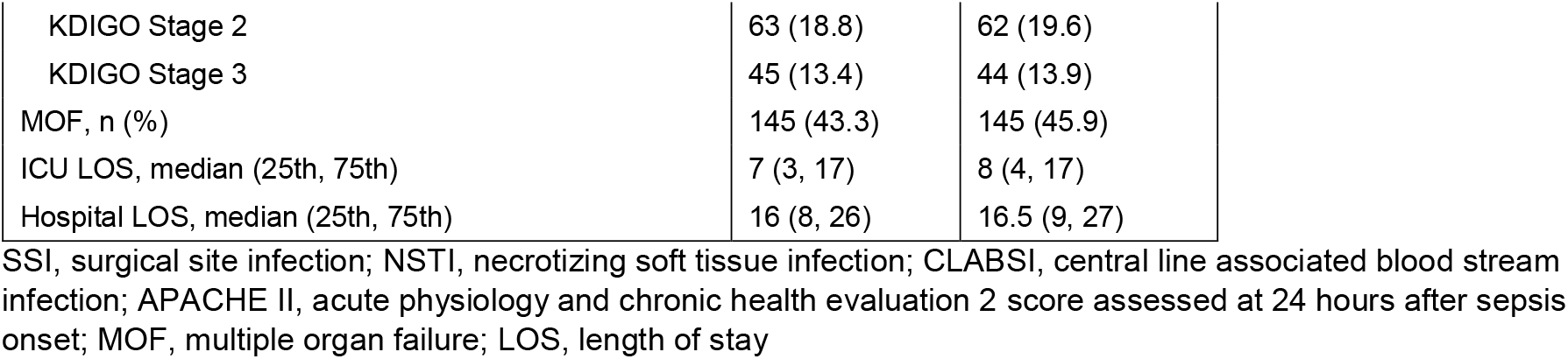
Cohort demographics.

**Supplemental Table 3.** Secondary Infections, Length of Hospitalization, and Discharge Disposition in the Overall and Sepsis-3 Cohorts. The two cohorts are further divided into subjects who experienced an early death, rapid recovery (RAP), or chronic critical illness (CCI).

| Outcomes | Overall cohort (n=335) | Early Death (n=11) | CCI (n=120) | RAP (n=204) | Sepsis -3 cohort (n=316) | Early Death (n=11) | CCI (n=119) | RAP (n=186) |
| --- | --- | --- | --- | --- | --- | --- | --- | --- |
| 2° infections/patient, mean (SD) | 0.5 (0.8) | 0.4 (0.5) | 1 (1) | 0.2 (0.5) | 0.5 (0.8) | 0.4 (0.5) | 1 (1) | 0.2 (0.5) |
| 2° infections/100 hospital days, mean (SD) | 2 (3.6) | 4.3 (7) | 3 (3.5) | 1.3 (3.2) | 2.1 (3.6) | 4.3 (7) | 3 (3.5) | 1.4 (3.3) |
| ICU LOS, median (25th, 75th) | 7 (3, 17) | 6 (5, 9) | 20 (15, 28.5) | 4 (2, 8) | 8 (4, 17) | 6 (5, 9) | 20 (15, 29) | 5 (3, 8) |
| Hospital LOS, median (25th, 75th) | 16 (8, 26) | 6 (5, 13) | 28 (20, 40) | 10.5 (7, 17.5) | 16.5 (9, 27) | 6 (5, 13) | 28 (20, 40) | 11 (8, 18) |
| Discharge disposition, n (%) |  |  |  |  |  |  |  |  |
| “Good” disposition | 193 (57.6) | 0 (0) | 27 (22.5) | 166 (81.4) | 175 (55.4) | 0 (0) | 27 (22.7) | 148 (79.6) |
| Home | 66 (19.7) | N/A | 1 (0.8) | 65 (31.9) | 58 (18.4) | 0 (0) | 1 (0.8) | 57 (30.6) |
| Home healthcare services | 102 (30.4) | N/A | 15 (12.5) | 87 (42.6) | 93 (29.4) | 0 (0) | 15 (12.6) | 78 (41.9) |
| Inpatient rehab facility | 25 (7.5) | N/A | 11 (9.2) | 14 (6.9) | 24 (7.6) | 0 (0) | 11 (9.2) | 13 (7) |
| “Poor” disposition | 142 (42.4) | 11 (100) | 93 (77.5) | 38 (18.6) | 141 (44.6) | 11 (100) | 92 (77.3) | 38 (20.4) |
| LTAC | 52 (15.5) | N/A | 48 (40) | 4 (2) | 51 (16.1) | 0 (0) | 47 (39.5) | 4 (2.2) |
| Skilled Nursing facility | 44 (13.1) | N/A | 11 (9.2) | 33 (16.2) | 44 (13.9) | 0 (0) | 11 (9.2) | 33 (17.7) |
| Another Hospital | 13 (3.9) | N/A | 12 (10) | 1 (0.5) | 13 (4.1) | 0 (0) | 12 (10.1) | 1 (0.5) |
| Hospice | 8 (2.4) | N/A | 7 (5.8) | 0 (0) | 8 (2.5) | 0 (0) | 7 (5.9) | 0 (0) |
| Death | 26 (7.8) | 11 (100) | 15 (12.5) | 0 (0) | 26 (8.2) | 11 (100) | 15 (12.6) | 0 (0) |

CCI, chronic critical illness; RAP, rapid recovery; LOS, length of stay; LTAC, long-term acute care facility. Early death, CCI and RAP percentages are those of their respective cohort (Overall, Sepsis-3).

**Supplemental Table 4.**
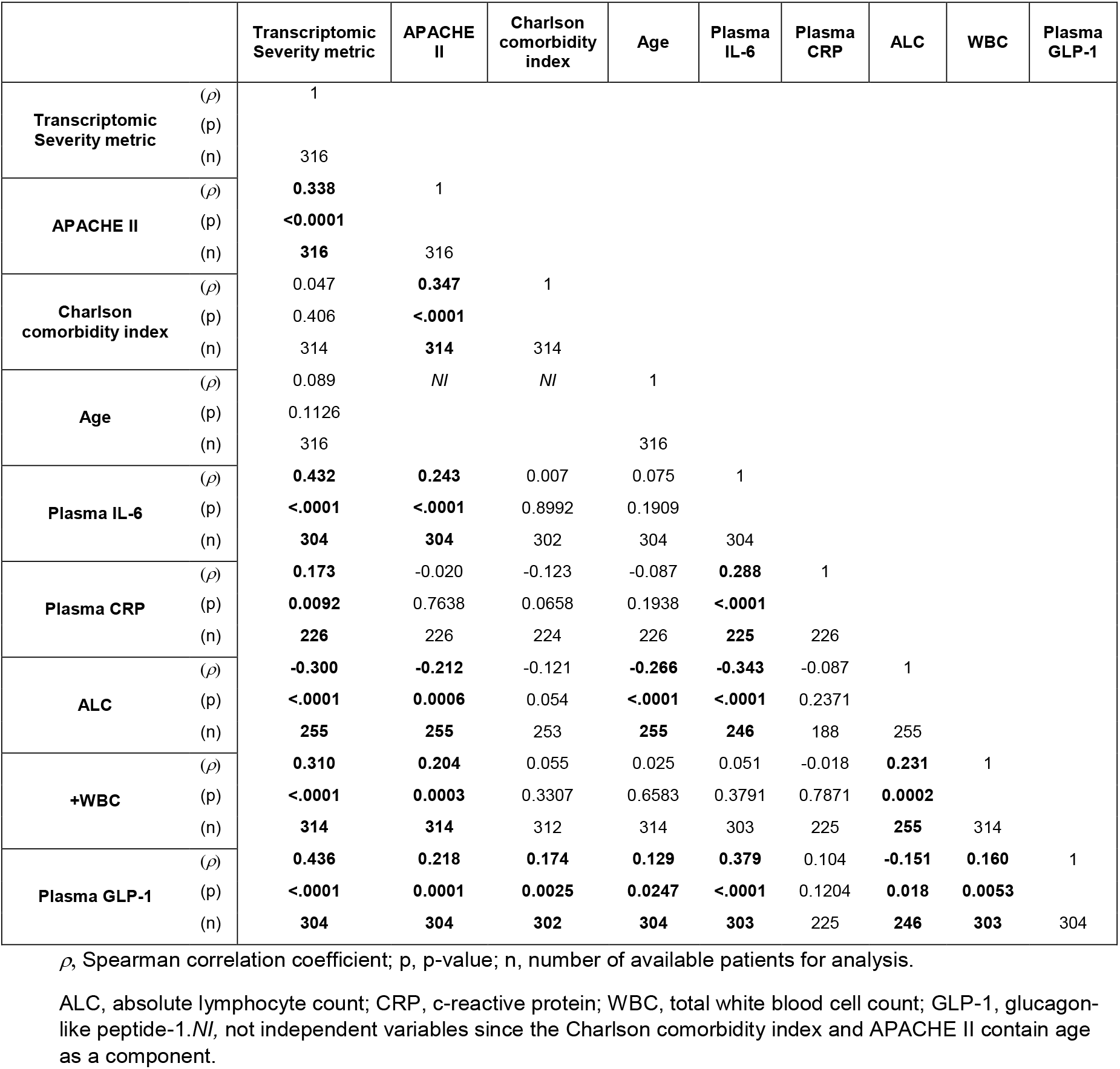
Correlation Coefficients of Biochemical and Clinical Markers obtained at 24 hours Post Sepsis. For patients meeting Sepsis-3 criteria, measurements were analyzed by Spearman’s correlation tests, as appropriate. Values present *rho* value, probability and sample size.

*Likelihood of a Viral Infection*. None of the patients were clinically adjudicated to have a viral infection. Routine viral DNA or antigen titers were not performed unless clinically justified, and none of the patients were considered to have a concurrent viral infection. Expectedly, the IMX-BVN-2 metric identified 333 of 335 subjects as being either ‘Unlikely’ or ‘Very Unlikely’. Only four subjects were deemed to have a ‘Possible’ or ‘Very Likely’ chance of having a viral infection (**Supplemental Table 5**). Since viral DNA or antigen titers were not conducted on any patient, it is impossible to determine whether a subclinical viral infection (or viral reactivation in the setting of critical illness) was present in these four patients with possible or very likely scores.

**Supplemental Table 5.**
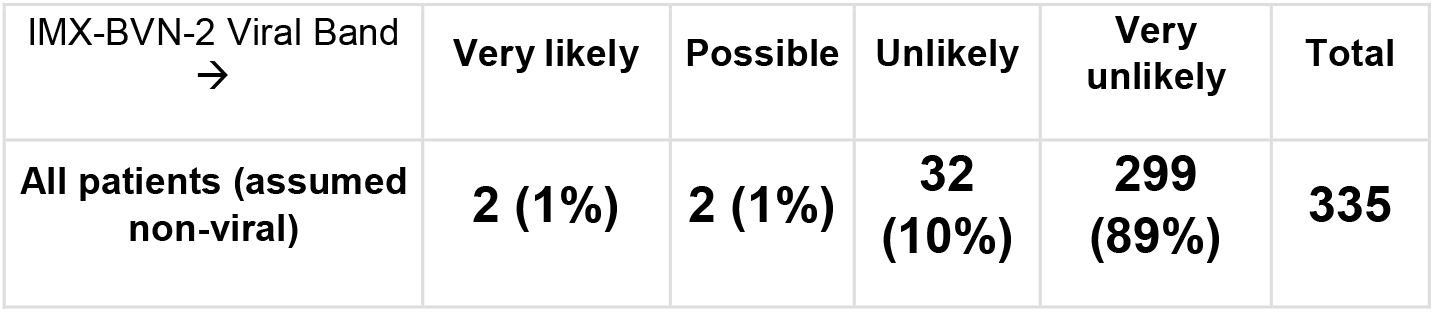
Distribution of IMX-BVN-2 Viral Infection Scores.

*Likelihood of a Bacterial Infection*. Importantly, this study was not designed to identify whether the metric could predict whether the patients had or did not have a bacterial infection (**Supplemental Table 6**). All of the patients were clinically adjudicated to have a microbial infection, either bacterial or fungal at the time of initiation of the sepsis management bundles. In addition, all of the patients had received broad spectrum antibiotics upon initiation of sepsis management bundles. In most cases, this was initiated 12-18 hours prior to blood sampling. At the time the study sample was collected, blood or exudate samples were not collected, so it would not be possible to determine how many of these subjects would still be infected, or the temporal dynamics of the IMX-BVN-2 score with regard to pre/post antibiotics timing.

**Supplemental Table 6.** Distribution of IMX-BVN-2 Bacterial Infection Scores.

| IMX-BVN-2 Bacterial Band<br>→ | Very likely | Possible | Unlikely | Very unlikely | Total |
| --- | --- | --- | --- | --- | --- |
| **Fungal infection** | **7 (78%)** | **2 (22%)** | **0 (0%)** | **0 (0%)** | **9** |
| **Bacterial infection** | **258 (79%)** | **63 (19%)** | **5 (2%)** | **0 (0%)** | **326** |

The IMX-BVN-2 metric identified both adjudicated fungal and bacterial infections as being ‘Very Likely’ or ‘Possible’ bacterial infections. Only 5 of the 335 included subjects were scored to be “Unlikely” whereas both the subjects with bacterial and fungal infections had close to 80% being very likely and nearly 20% as possible.

## Notes

Conflicts of Interest and Source of Funding: This project has been funded in part with U.S. Federal funds from the Department of Health and Human Services; Office of the Assistant Secretary for Preparedness and Response; Biomedical Advanced Research and Development Authority (BARDA), DRIVe program, under Contract 75A50119C00044. The original observational clinical study was funded under P50 GM-111152, awarded by the National Institute of General Medical Sciences (NIGMS). DD and BF were funded by a post-graduate training grant in burns, trauma, sepsis and perioperative injury from the NIGMS (T32 GM-008721). UM, JW, PL, OL and TE are employees of Inflammatix, Inc. (Burlingame, CA, USA). SB, PS, and LLM received research support for aspects of this study under BARDA subcontract. The remaining authors have disclosed that they do not have any conflicts of interest.

### Clinical Trial

NCT02276417

### Funding Statement

This project has been funded in part with U.S. Federal funds from the Department of Health and Human Services; Office of the Assistant Secretary for Preparedness and Response; Biomedical Advanced Research and Development Authority (BARDA), DRIVe program, under Contract 75A50119C00044. The original observational clinical study was funded under P50 GM-111152, awarded by the National Institute of General Medical Sciences (NIGMS). DD and BF were funded by a post-graduate training grant in burns, trauma, sepsis and perioperative injury from the NIGMS (T32 GM-008721).

### Author Declarations

Ethics approval was obtained from the University of Florida Institutional Review Board (#201400611).

